# DEVELOPMENT AND TEST OF A CHATBOT TO IDENTIFY DEPRESSION

**DOI:** 10.1101/2021.10.28.21265110

**Authors:** Stefano Neto Jai Hyun Choi, Rita Simone Lopes Moreira, Ana Luiza Fontes de Azevedo Costa, Caio Vinicius Saito Regatieri, Vagner Rogério dos Santos

## Abstract

**Purpose:** to develop and test a prototype of Chatbot with the purpose of applying a questionnaire to assess depression in visually impaired individuals.

**Methods:** This project was carried out in the Federal University of São Paulo. The Chatbot was developed using the platform BLiP®. The social-demography questionnaire and the Center for Epidemiological Scale – Depression (CES-D) were selected to collect the essential data and to identify the presence of depression, respectively. After the development, validation tests were applied to verify the functionality and structure of the chatbot.

**Results:** The Chatbot prototype presented an excellent flow of conversation in the tests conducted. The questionnaires were applied in a satisfactory manner during the tests. Software validation tests approved the prototype’s function.

**Conclusions:** The Chatbot prototype is an affordable and easy way to apply questionnaires that can be used to identify health conditions, such as the likelihood of being depressed.

## INTRODUCTION

The most recent data collected by the Vision Loss Expert Group showed that 441,1 million people are visually impaired worldwide. Of these, 36 million are legally blind. This represents a prevalence of visual impairment of 5.52% and 0.49% of legal blindness in the global population. ^1^

The visual impairment is defined by the International Classification of Diseases 11 (2018) as distance or near vision impairment. The distance vision impairment is divided in mild (visual acuity worse than 6/12 to 6/18), moderate (worse than 6/18 to 6/60), severe (worse than 6/60 to 3/60) and blindness (worse than 3/60). The near vision impairment is visual acuity worse than N6 or M.08 at 40cm. ^2^

The visual acuity (VA) is the measurement of the sharpness of vision. This measurement defines how the eye discerns shapes, letters or numbers on specific charts. The VA can be measured monocularly and/or binocularly. ^3^

In 2010, the Brazilian Geography and Statistic Institute (IBGE) found that 45 million Brazilians had some type of impairment, being 35.7 million visual impairment. ^4^

Despite the interventions available, a study done by Ferracina et al. demonstrated that anxiety and tendency to depression are common in adult patients with glaucoma. ^5^ In 2006, another study showed that patients with uveal melanoma in different treatment stages presented depression requiring psychological monitoring. ^6^

Depression is a mood disorder caused by many factors, conditions, events and/or disorders that can affect any individual. ^7^

Depression associated with ocular disorders is a topic that has been discussed by specialists in medicine. In North America, 22% to 38% of patients attending visual rehabilitation centers presented some degree of depression. ^8^

According to Adamson, patients that suffered from untreated depression associated with visual impairment had a worse quality of life. ^9^ The association of depression and visual impairment can compromise the general state of health, worsen the physical and mental condition, decrease the activity of the economically active population, and decrease the quality of life overall. ^10^ Major depression is among the lead causes of loss of years of productive life. ^11^

In 2016, the World Bank and the World Health Organization (WHO) emphasized the necessity to invest in mental health treatment, since 350 million people suffered from depression during that year. In Brazil, it is estimated that 10% to 18% of the population suffered from depression in the same year. Therefore, around 10% of the people with depression in the world were located in Brazil in 2016. ^12^

A study made by Chisholm et al. revealed that the cost of extending and amplifying the antidepressant treatment would be about 137 billion dollars, while properly treating more patients would yield a profit of about 300 billion dollars more than expected to economy. ^13^

The total of professionals that diagnose and treat mental disorders is still low. In developed countries, there are only nine psychiatrists for 100,000 citizens. In developing countries, there is only 0.01 psychiatrist for 100,000 citizens. ^11^

The CES-D is a great instrument when considering the possibility of using Artificial Intelligence (AI) in the pre-diagnosis of depressive states. This questionnaire does not require the administration by a trained professional, and it is considered gold standard to identify depression. Furthermore, a study published in 2010 validated and translated it to Portuguese. ^14^

Considering all the risks related to depression and the scarcity of psychiatrists, Virtual Assistants were developed based on artificial intelligence (AI) to prevent suicide and administrate Cognitive Behavioral Therapy (CBT). The first example is Ellie, which is a chatbot that administrates a phycological test with American soldiers to identify symptoms of Post-Traumatic Stress Disorder (PTSD). Another example of a chatbot is Sara. Sara offers mental healthcare by daily texting patients to keep track of them. ^11^

Chatbots (CB) are based in Artificial Intelligence and are developed to stimulate a conversation between a person and the virtual assistant, offering questions and answers by the use of Natural Language Processing (NLP). NLP is an instrument of AI labored to comprehend and to answer assertively. It can adapt and learn whenever necessary, making the user feel like he/she is interacting with another person, not only a virtual assistant. ^15^

The benefits of using CB in medicine are the possibility of 24 hours monitoring, personalized assessment, decrease in waiting time in queues, prevention of unnecessary visits to the hospital, reduction in costs, and ability to answer FAQ (Frequent Asked Questions). ^16, 17^

The CB development was done in two phases. The first one was the development of a Specialist System (SS) based on the Sociodemographic questionnaire and the CES-D questionnaire. The second phase was the validation of the prototype. ^18^

## METHODS

This study was analyzed and approved by the Research Ethics Committee of UNIFESP/HSP under the number 2097051017.

To develop the SS in order to apply a questionnaire to assess the presence of depression (CES-D), the knowledge base used was constituted of three instruments, the Sociodemographic Questionnaire, the CES-D, and a Platform to create chatbots named BLiP®. In this platform. ^19^

Features as age, financial condition and gender can be related to the cause of depression. The sociodemographic questionnaire used by the Psychobiology Department of UNIFESP was selected to collect all of the necessary data from the users.

The questionnaire used for depression assessment CES-D was introduced in the prototype.

In BLiP®, the two selected questionnaires were inserted in the prototype. Facebook Messenger® was chosen to be the test environment.

### Patient Entry

The process starts when the user is not feeling well and contacts Julie2019, the prototype developed in this study.

As shown in figure 1, it is possible to identify the name of the Facebook Messenger’s user since the first message. It evidences the proposed benefit that is the attempt to keep a human conversation personalized and assertive.

**Figure 1.**
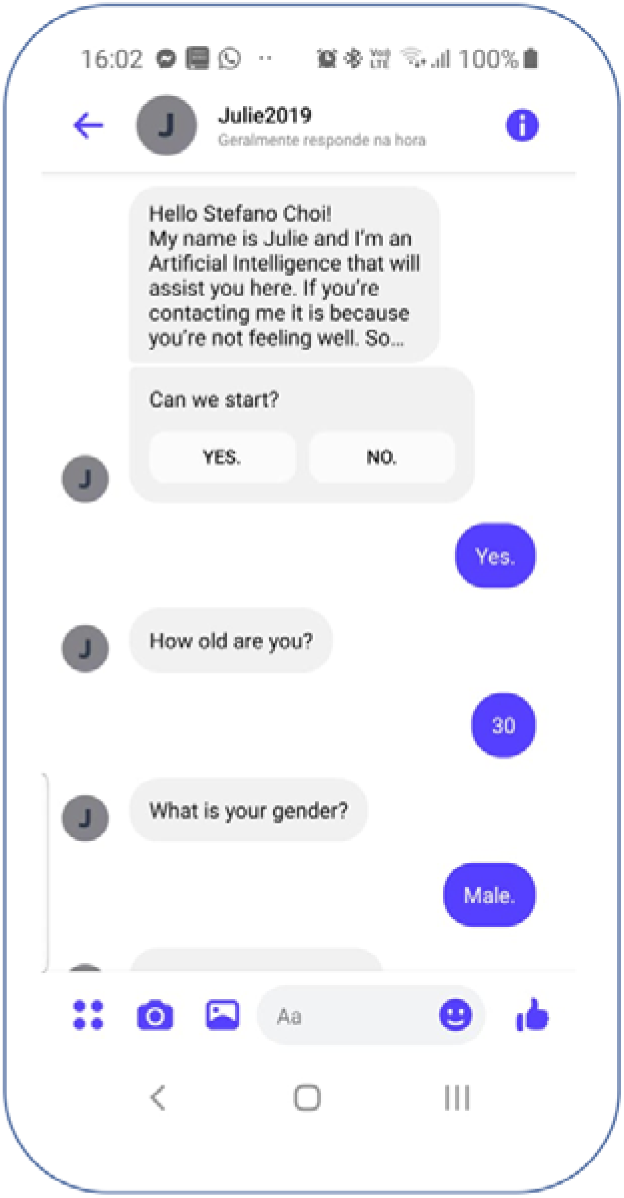
First Message Sent by the prototype. It identifies the name of the user of the Messenger®, customizing the conversation. After the introduction, the prototype will ask the user for permission to start the dialogue. The authors took a print screen from the smartphone used on the tests.

The prototype must identify itself as a virtual assistant to prevent confusion and avoid any emotional bond or affection. For example, Xiaoce®, a chatbot developed to help and comfort people that suffered love deception, confused its users to the point that they believed that it was human and even fell in love with it. ^20^

### Permission to Administrate the Sociodemographic Questionnaire

Initially, a message is sent to the user asking permission to start the questions. This type of interaction is meant to stimulate the user to be more interactive and to pay more attention to the next questions.

### Sociodemographic Questionnaire

The next protocol step is the sociodemographic questionnaire administration. The collection of this data has been shown to be important in depression assessment questionnaires already validated in Brazil, and is useful to better direct the Chatbot interaction with the user. Figure 2 presents the structure of the Sociodemographic questionnaire on Messenger®.

**Figure 2.**
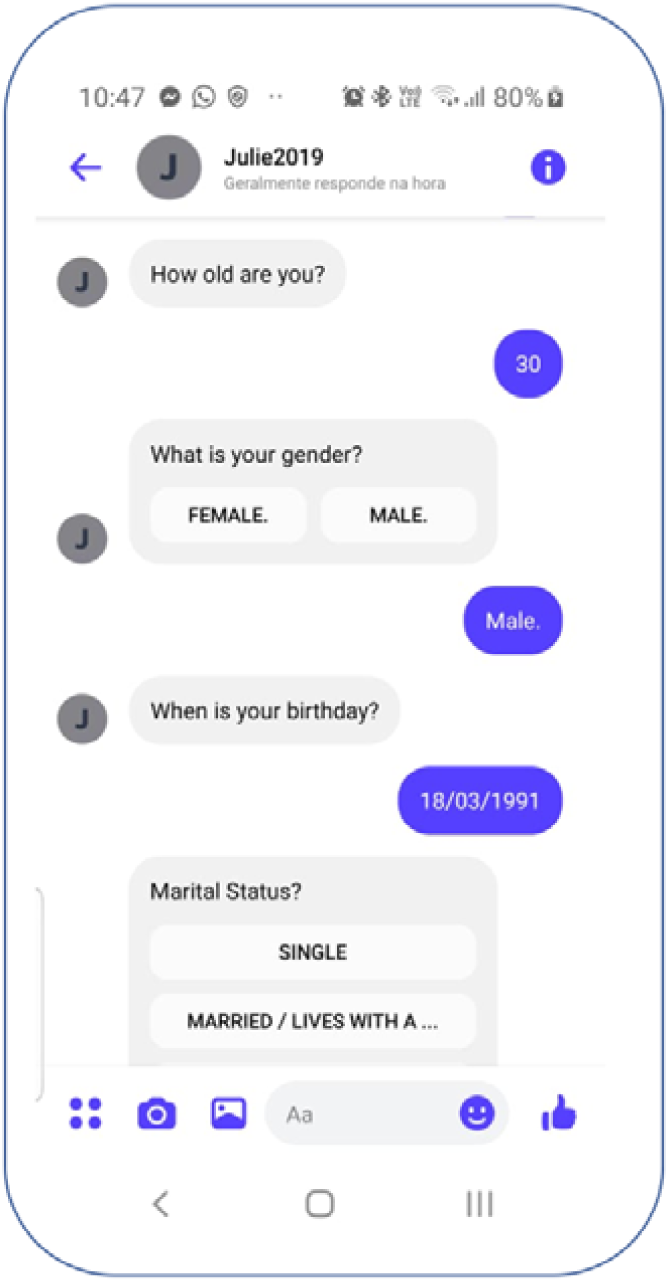
Sociodemographic Questionnaire on Messenger®. The authors took a print screen from the smartphone used on the tests.

### Notification of the CES-D Questionnaire Initiation

The prototype informs the user that the questionnaire requires between fifteen to thirty minutes without interruptions to yield a reliable result and to avoid possible bias.

At the end of the questionnaire, the prototype will add up the scores of each question. If the sum is between 0 to 11 points, depression is unlikely, but if it is between 12 to 60 points, depression is more likely present.

### Validation Test

Firstly, this prototype was tested using a Structural test, known as the White Box Test. This test evaluates the internal behavior of the software. The developer accessed the development platform and verified each entry and exit conditions of all message boxes in the prototype. Conversation Tests flow and variations of possible user’s answers were applied. ^21^

The Functional Test, or Black Box Test, was conducted after the White Box Test. The developer became a user and interacted with the prototype several times to verify the expected answers without accessing the internal structure of the prototype. ^21^

Finally, a Validation Test was performed to compare the results obtained randomly from the CES-D with the results obtained by the prototype. For each of the 20 questions on the CES-D, one of the answers was randomly selected from the 4 possible ones. The random selection was performed based on the fact that all the questions on the CES-D are equally important, which means that the only criteria that defines the likelihood of depression is the sum of the score for each answer.

Six CES-D questionnaires were randomly answered and compared to those answered in the prototype, to verify and validate the potential use of the prototype in applying the CES-D questionnaire.

## RESULTS AND DISCUSSION

In the White Box Test, the developer assessed the structure and the code of the prototype inside the BLiP® platform. All the stages of the protype construction were verified to correct and repair any error found. The second test consisted of several interactions between the prototype and the developer as a user. In both, the developer didn’t find any issues or errors. Nevertheless, all the software has to be revised and tested periodically. ^21^

Six CES-D questionnaires were manually filled by randomly selecting one of the answers to each question. The final score of each questionnaire defines the likelihood of being depressed, and these results were used as a reference to test the prototype. The randomization was useful to demonstrate that, regardless of the answers provided, whenever applied by the prototype, the same score will be obtained, making it possible to compare and asses the prototype functionality.

The comparison showed that the questionnaire applied by the chatbot performed as well as the manual one. This way, we verified that the prototype could serve as a virtual assistant to apply the CES-D questionnaire.

For the prototype to be an accessible tool to visually impaired individuals, it is necessary to adapt the size of the font on the smartphone according to the visual acuity of each user. In this study, the Samsung® Galaxy s8 was used.

A study conducted by Bailey in 1998 developed a table to relate the visual acuity with the font size that would be ideal to enable reading at 40cm, which is a distance considered comfortable for reading. Another study from 2019 presented the ideal printed font sizes that could be read according to the visual acuity. The table 1 shows the relationship between visual acuity, ideal font size and ideal printed font size to be read at 40 cm. ^22, 23^

**Table 1.** Printed font Size Correlated with Visual Acuity from a Distance of 40cm.

| **Visual Acuity (Snellen)** | **Notation M** | **Size – Points (font)** | **Font size in millimeters/height X** | **Printed font size in millimeters** |
| --- | --- | --- | --- | --- |
| 20/20 | 0.4 | 3.2 | 0.58 | 1.12 |
| 20/25 | 0.5 | 4 | 0.73 | 1.4 |
| 20/32 | 0.6 | 5 | 0.92 | 1.75 |
| 20/40 | 0.8 | 6 | 1.16 | 2.1 |
| 20/50 | 1 | 8 | 1.45 | 2.8 |
| 20/63 | 1.2 | 10 | 1.82 | 3.5 |
| 20/80 | 1.6 | 14 | 2.33 | 4.9 |
| 20/100 | 2 | 16 | 2.91 | 5.6 |
| 20/125 | 2.5 | 20 | 3.94 | 7 |
| 20/160 | 3 | 24 | 4.65 | 8.4 |
| 20/200 | 4 | 33 | 5.82 | 11.55 |
| 20/400 | 8 | 66 | 11.6 | 23.1 |

*This table presents the correlation between printed font size with visual acuity from a distance of 40cm.<sup>22, 23</sup>*

To change the size of the font in the smartphone used in this study, it was necessary to access the following: Configurations, Accessibility, Visibility Improvements, Font size and style and Adjust to maximum size.

After the configuration adjusts, the font size achieved was a minimum of 4 mm and maximum of 5 mm, which was not considered sufficient to reach the ideal printed font size recommended for visually impaired individuals M1.6 or worse. Therefore, it was necessary to utilize other tools from the smartphone to achieve larger font sizes. The sequence details the commands used: Configurations, Accessibility, Visibility Improvements and Amplification window.

This enables further enlargement of the font so that the prototype can be used by individuals with M1.6 near vision.

Figure 3 shows the smartphone font adjusted up to 30mm of height. This enables the use of the prototype by visually impaired individuals with varying degrees of visual acuity. Figure 4 presents the actual font size.

**Figure 3.**
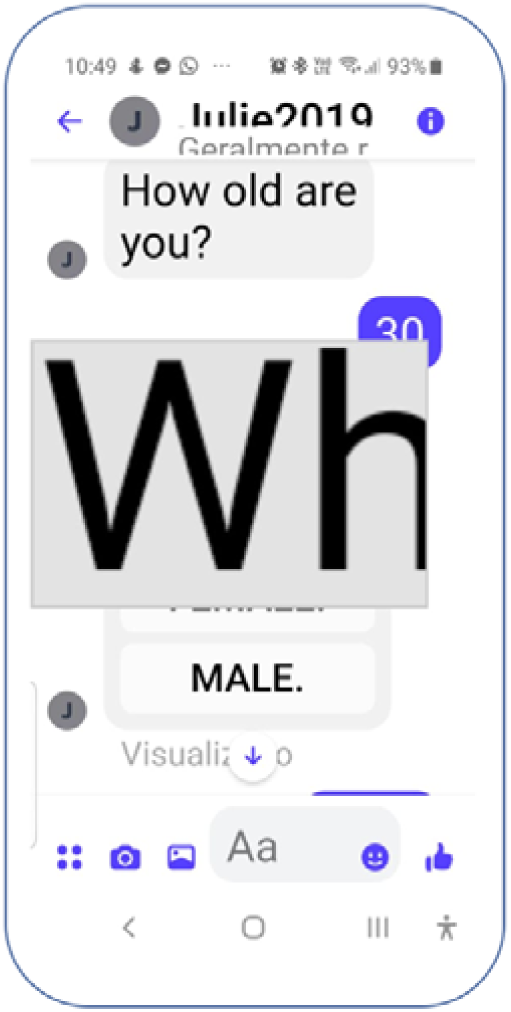
Smartphone font adjusted up to 30mm of height. The authors took a print screen from the smartphone used on the tests.

**Figure 4.**
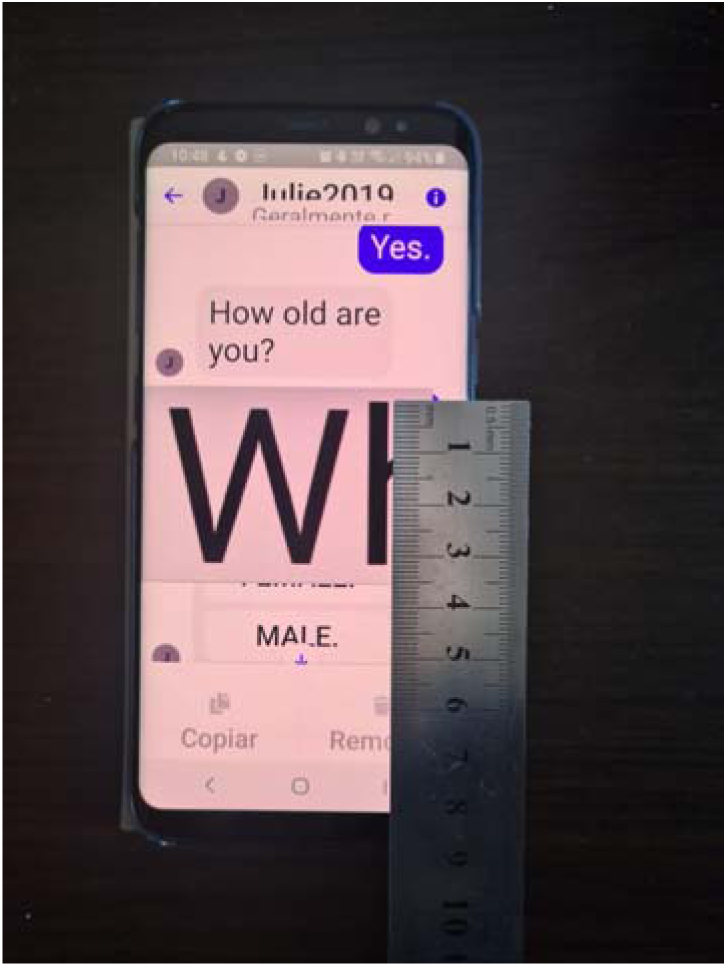
Actual font size on the smartphone.

The application of any questionnaires using CB doesn’t generate a diagnosis by this virtual assistant. Therefore, this prototype doesn’t provide a diagnosis, but identifies the likelihood of being currently depressed.

The results showed that the conversation flow didn’t present any issues. However, a possible problem that could cause confusion is when one question is sub-divided into more than three questions. In this case, the question was divided into two parts, as shown in figure 4. If the patient doesn’t perceive this separation, he/she may answer incorrectly. The prototype can guide the patient to pay attention when answering to avoid this kind of bias.

One of the advantages of the Chatbot is the possible application of AI. The prototype developed in this project does not have AI implemented because both questionnaires had predefined answers. The SS doesn’t need to understand the language to answer correctly, and the order of the questions was respected.

Although the chatbot was programmed to be used with humans undergoing ophthalmic treatment, we initially validated the software to assess the ability to apply the CES-D questionnaire. The next phase of this research will be the test of the Chatbot prototype with humans.

## CONCLUSION

From the results, we concluded that the Chatbot can be a useful tool to aid in the assessment of symptoms through questionnaires. The technology innovations available are not meant to substitute the health care professionals, but instead to be a tool to improve health care services by reaching remote areas and identifying individuals that may be in need of a timely intervention.

## Data Availability

All data produced in the present work are contained in the manuscript

